# Comparative accuracy of five screening tools for sarcopenia in community older adults:a systematic review and a network meta-analysis

**DOI:** 10.1101/2024.04.16.24305890

**Authors:** Jie Li, Yujie Yang, Menglin Gao, Huaihong Yuan

**Affiliations:** Department of Nephrology, Kidney Research Institute, West China, Hospital of Sichuan University, Chengdu 610041, China; West China School of Nursing, Sichuan University, Chengdu 610041, China

**Author notes:** Corresponding authors: Huaihong Yuan Email addresses.

**Keywords:** community, older adults, sarcopenia, sensitivity, specificity, network Meta-analysis

## Abstract

**Background:** Sarcopenia, a prevalent and serious condition among community older adults, often remains unnoticed. The use of systematic screening has the potential to enhance detection rates; however, there is currently no consensus on the most effective approach. This study aimed to assess the diagnostic test accuracy of five simple sarcopenia screening tools and determine which test has the highest accuracy.

**Objective:** To assess and compare the accuracy of five screening tools for sarcopenia in community older adults.

**Design:** A systematic review and a network meta analysis.

**Methods:** A systematic search was conducted in various databases including Pubmed, The Cochrane Library, Embase, Web of Science, CNKI, Wanfang, VIP, and Sinomed up to September 2023. Studies reporting on the accuracy of diagnostic testing for sarcopenia in community-dwelling older adults using one or more of the following sarcopenia screening tools were included: Sarcopenia Simple Five-Item Rating Scale (SARC-F), SARC-F combined with calf circumference (SARC-CalF), SARC-F combined with older adults and BMI (SARC-F+ EBM), Mini sarcopenia risk assessment-5 (MSRA-5), and Mini sarcopenia risk assessment-7 (MSRA-7). The reference standard was the Asian Working Group on Sarcopenia (AWGS), the European Working Group on Sarcopenia on Older People (EWGSOP), the Foundation for National Institutes of Health (FNIH), or the International Working Group on Sarcopenia (IWGS). Random-effects bivariate binomial model meta-analyses, meta-regressions and a network meta-analysis were used to estimate the pooled and relative sensitivities and specificities.

**Results:** We identified and evaluated 22 papers focused on SARC-F, S ARC-CalF, MSRA-5, and MSRA-7. Traditional meta-analysis sorting results showed summary sensitivities of 0.25, 0.59, 0.43, 0.82, and 0.51, summary specificities of 0.94, 0.82, 0.81, 0.39, and 0.85, summary AUC of 0.80, 0.76, 0.70, 0.68, and 0.75, and summary DOR of 5, 7, 3, 3, and 6. The network meta-analysis ranking results showed that MRSA-5 had the highest sensitivity (92.27) and SARC-F had the highest specificity (99.81) under the cumulative ranking.

**Linking evidence to action:** The MSRA can be used as a tool for screening sarcopenia in community older adults, while the SARC-F can be used for first-time diagnosis of sarcopenia in this population. However, it is important to interpret the results with caution due to the variability among different studies analyzing the accuracy of this diagnostic test. Future research should focus on obtaining additional evidence from large sample sizes and high-quality studies.

## 1. Introduction

Rosenberg firstly introduced the concept of “sarcopenia” in 198 9 and described it as an age-related loss of muscle mass, muscle strength and muscle function (Rosenberg, 1997). This condition is associated with adverse outcomes such as falls, functional decline, frailty, and even death (Cruz-Jentoft and Sayer, 2019). In October 2016, the World Health Organization officially recognized sarcopenia as an independent clinical condition and included it in the International Classification of Diseases ICD-10 codes (Anker et al., 2016). According to statistics, the number of people suffering from sarcopenia is currently as high as 50 million worldwide (Yuan and Larsson, 2023), with a prevalence of sarcopenia of around 10-27 percent among healthy older people aged 60 years and above (Petermann-Rocha et al., 2 022). Therefore, there is significant clinical importance in rapidly screening, identifying, and intervening for sarcopenia among older adults in the community.

Currently, there is no standardized diagnostic criteria for sarcopenia. Various diagnostic criteria have been proposed by different organizations such as the Asian Working Group on Sarcopenia (AWG S) (Chen et al., 2020), the European Working Group on Sarcopenia on Older People (EWGSOP) (Cruzjentoft et al., 2010), the Foundation for the National Institutes of Health (FNIH) (Lee and David, 2010), and the International Working Group on Sarcopenia (IWGS) (Cesari et al., 2012). However, these criteria involve complex operations and the use of advanced equipment like CT scans, MRI, X-ray bon e densitometry, and bioelectrical impedance analyzers. As a result, they are not suitable for early identification of sarcopenia or large-scale screening. It is important to develop a simple yet effective screening tool that can enable caregivers to detect sarcopenia in older individuals at an early stage. This would facilitate early diagnosis and treatment, preventing preclinical changes in sarcopenia and reducing the occurrence of negative consequences.

However, there are more than 10 clinically applied screening tools for sarcopenia with variable accuracy, and healthcare professionals are troubled by their selection. How to recommend appropriate screening tools to efficiently identify the risk of sarcopenia in the elderly is a pressing issue in clinical practice. Commonly used screening tools for sarcopenia include the Sarcopenia Five-Item Scale (SARC-F) (Malmstrom and Morley, 2013), the Sarcopenia Five-Item Combined Calf Circumference Scale (SARC-Calf) (Barbosa-Silva et al., 2016), and the Sarcopenia Five-Item Combined Aging and Body M ass Index Scale (SARC-F+EBM) (Kurita et al., 2019). Additionally, the Mini Sarcopenia Risk Assessment (Rossi et al., 2017) offers a 5-item questionnaire (MSRA-5) or a 7-item questionnaire (MSRA-7). An ideal sarcopenia screening test should have high sensitivity and specificity, with a threshold of over 80% to ensure effectiveness. By comparing the effectiveness of individual tests, we can determine their relative diagnostic test accuracy. These comparisons can provide valuable insights for clinicians and researchers in selecting the most suitable tests for their specific purposes. It is important to note that no previous systematic reviews comparing the test accuracy of common sarcopenia screening tools were found in our research.

To address this research gap, we performed a comprehensive systematic review and network meta-analysis. Our aim was to identify and compare the diagnostic test accuracy of commonly used sarcopenia screening tools in community-dwelling older adults. We specifically evaluated these tools against internationally recognized diagnostic criteria for sarcopenia.

## 2. Materials and methods

A protocol for this review was registered in PROSPERO **(CRD 42023487209).** The conduct and reporting of this systematic review adhered to the guidelines specified in the Cochrane Handbook for Systematic Reviews of Diagnostic Test Accuracy and the Preferred Reporting Items for Systematic Reviews and Meta-Analyses (PRISMA 2020 Statement) (Page et al., 2021). Additionally, we incorporated pertinent components from the Guidelines for Reporting Systematic Reviews in Network Meta-Analyses (PRISMA-NMA) (Hutton et al., 20 16) and the Guidelines for Reporting Methodological Quality of Systematic Reviews (AMSTAR-2) (Shea et al., 2017).

### 2.1. Search strategy

We conducted a comprehensive search of electronic databases, including Pubmed, the Cochrane Library, Embase, Web of Science, CNKI, Wanfang Data, VIP, and Sinomed. Each database was searched from inception to September 30, 2023. The article is based on all reported studies on screening for sarcopenia in community-dwelling older adults. All database searches were conducted using a combi nation of subject terms and free words. The search terms used were: (aged OR elder OR old people) AND (sarcopenia* OR muscle contraction OR muscle atrophy OR SARC-F OR SARC-Calf OR SARC-F+EBM OR MSRA OR Sarcopenia Risk assessment OR Sarcopenia assessment tool OR Screening for Sarcopenia).

### 2.2. Inclusion criteria and exclusion criteria

The inclusion criteria for this study were as follows: (i) the study population consisted of older adults in the community; (ii) the study was a diagnostic study published in English or Chinese; (iii) the diagnostic methods included five commonly used community sarcopenia screening scales, namely the Sarcopenia Five-Item Scale (S ARC-F), the Sarcopenia Five-Item Combined Calf Circumference Scale (SARC-Calf), and the Sarcopenia Five-Item Combined Aging an d Body Mass Index Scale (SARC-F+EBM). Additionally, the Mini Sarcopenia Risk Assessment offers a 5-item questionnaire (MSRA-5) or a 7-item questionnaire (MSRA-7); (iv) the diagnostic gold standard used was AWGS, EWGSOP, FNIH, or IWGS; (v) the outcome indicators of interest were sensitivity or specificity.

The exclusion criteria were as follows: (i) literature with incomplete data extraction; (ii) conference abstracts, reviews, dissertations, and other similar publications; (iii) duplicate publications.

### 2.3. Scales

The SARC-F was compiled by Malmstrom (Malmstrom and Morley, 2013) in 2013. It is mainly used for community elderly and hospitalized elderly patients. It is the most widely used tool for screening sarcopenia in the elderly. The questionnaire needs to be completed under the guidance of medical staff and includes 5 aspects: muscle strength, assisted walking, sitting up, climbing stairs and number of falls. Each item is scored from 0 to 2 points, and the total score is from 0 to 10 points. If the score is ≥ 4 points, sarcopenia can be initially suspected.

The SARC-CalF adds the calf circumference item compared to SARC-F. It was originally invented by Brazilian scholar Barbosa-Silva (Barbosa-Silva et al., 2016) and others, and the evaluation met hod is the same as SARC-F.

Kurita (Kurita et al., 2019) developed the SARC-F+EBM, which incorporates the concepts of ‘E’ (elderly) and ‘BMI’ (body mass index) into the original SARC-F questionnaire. In addition to the SARC-F score, this method takes into consideration whether the patient is 75 years old or above, and whether their BMI is ≤ 21 kg/m2. For patients below 75 years old, the score remains at 0 points. However, for patients aged 75 years or older, the score is increased to 10 points. Similarly, if the BMI is ≤ 21 kg/m2, the score remains at 0 points, but if the BMI is >21 kg/m2, the score is increased to 10 points. The total score ranges from 0 to 30 points, and a score of ≥ 12 points indicates a positive result. The evaluation method remains the same as for the original SARC-F.

The Mini Sarcopenia Risk Assessment Questionnaire, developed by Rossi (Rossi et al., 2017) in 2017, consists of two versions: MSRA-5 and MSRA-7. MSRA-7 includes factors such as age, physical activity level, number of hospitalizations in the previous year, weight loss, regularity of three meals, dairy intake, and protein intake. A score of ≤ 30 on MSRA-7 indicates a risk of sarcopenia. On the other hand, MSRA-5 does not include two items: regular meals and dairy intake. A score of ≤ 45 on MSRA-5 indicates a risk of sarcopenia. It is important to note that completing this questionnaire requires the guidance and assistance of medical staff.

### 2.4. Literature screening and data extraction

The assessment of literature quality and data extraction were conducted independently by two researchers. Any disagreements were resolved through mutual discussion or arbitration by a third researcher. Data extraction of the included literature was performed after initial screening and re-screening using the literature management software EndNote X9. The extracted data included information such as the first author, year, country, sample size, age, screening tool, reference gold standard, true positive, false positive, true negative, false negative, sensitivity, and specificity.

### 2.5. Quality assessment

The literature was assessed for quality using the Quality Assessment Tool for Diagnostic Accuracy Studies (QUADAS-2) (Whiting et al., 2011) by two independent researchers. Disagreements between the researchers were resolved through discussion. The tool evaluates both the risk of bias and clinical applicability.

### 2.5. Statistical methods

The traditional meta-analysis was conducted using Stata (version 17.0). Sensitivity, specificity, positive likelihood ratio, negative likelihood ratio, diagnostic odds ratio (DOR), and 95% confidence interval (CI) were pooled for each literature. Network evidence was visualized, consistency tests were performed, and funnel plots were created using the midas command in the metan module.

The Bayesian network Meta-analysis was conducted using the gemtc code package of R (version 4.2.1). The analysis involved 4 chains, 100,000 iterations, and a step size of 10. The area under the cumulative ranking probability curve (SUCRA) was used to determine the best screening tool. The SUCRA value indicated the likelihood of each screening tool being the best. The sensitivity and specificity of the screening tools were ranked based on the SUCRA value. The odds ratio (OR) and its 95% confidence interval (CI) were calculated, and statistical significance was determined when the 95% CI of the OR value did not include 1.

## 3. Results

### 3.1. Search results

Fig. 1 shows the details of the study selection process. Out of the initial 3293 records, we identified 2782 unique studies after removing duplicate publications. After a preliminary screening of titles and abstracts, we found 142 articles that were potentially relevant to the use of the five scale for screening sarcopenia in the elderly. Following a thorough full-text review, we excluded 120 studies, leaving us with 22 articles that met our eligibility criteria. It is worth noting that none of the 22 articles included in our analysis were from unpublished sources.

**FIGURE 1.**
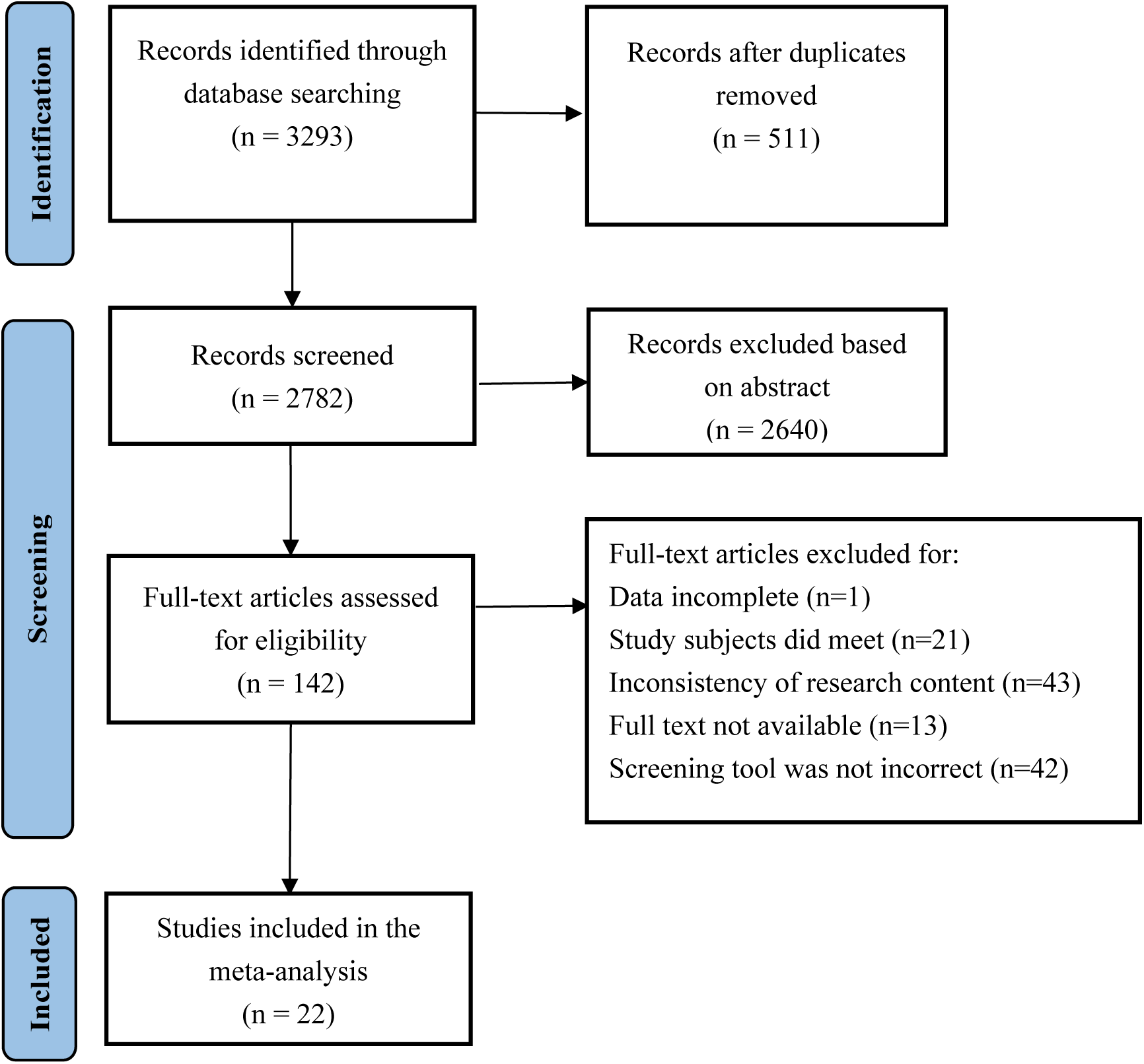
The flow chart of the search for eligible studies.

### 3.2. Study Characteristics

The analysis included a total of 22 studies with a combined sample size of 15,493 research subjects. All studies were published within the past five years, and the research areas involved China, Japan, Italy, Poland, South Korea, Brazil, Singapore, Japan, and Turkey. The age of patients included in the diagnostic test accuracy studies ranged from 60 to 94 years. The study sample sizes ranged from 73 to 4000. All sarcopenia screening tests are performed in community settings. Additionally, these studies utilized 5 different screening tools and each study was compared directly with the gold standard. Out of the 22 studies, 13 of them compared two or more tools with the gold standard, but there were no direct comparisons between two tools, so separate sets of data were extracted. The most commonly used sarcopenia screening tool was SARC-F. Table 1 presents the characteristics of the 22 included studies.

**TABLE 1.**
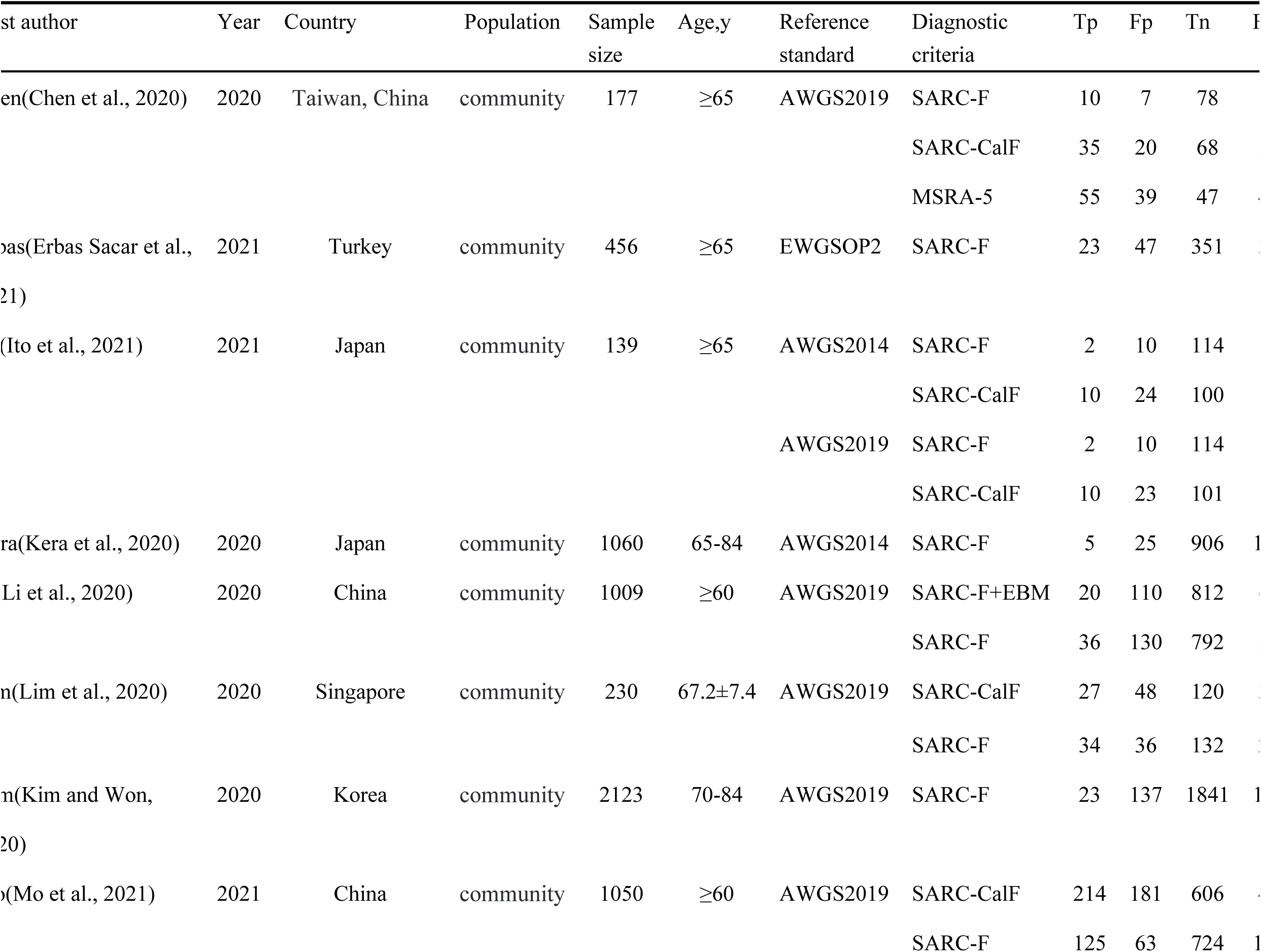
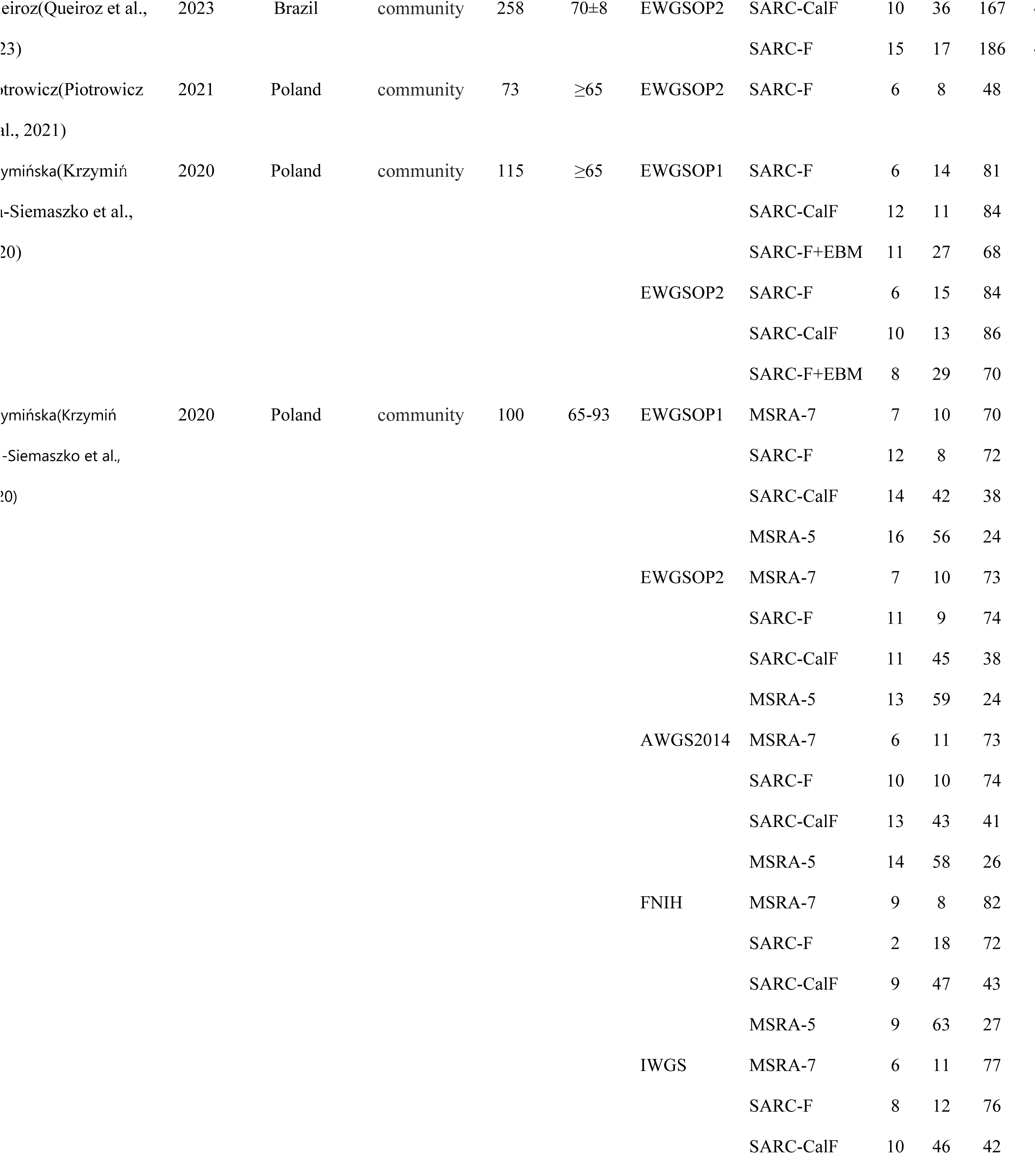
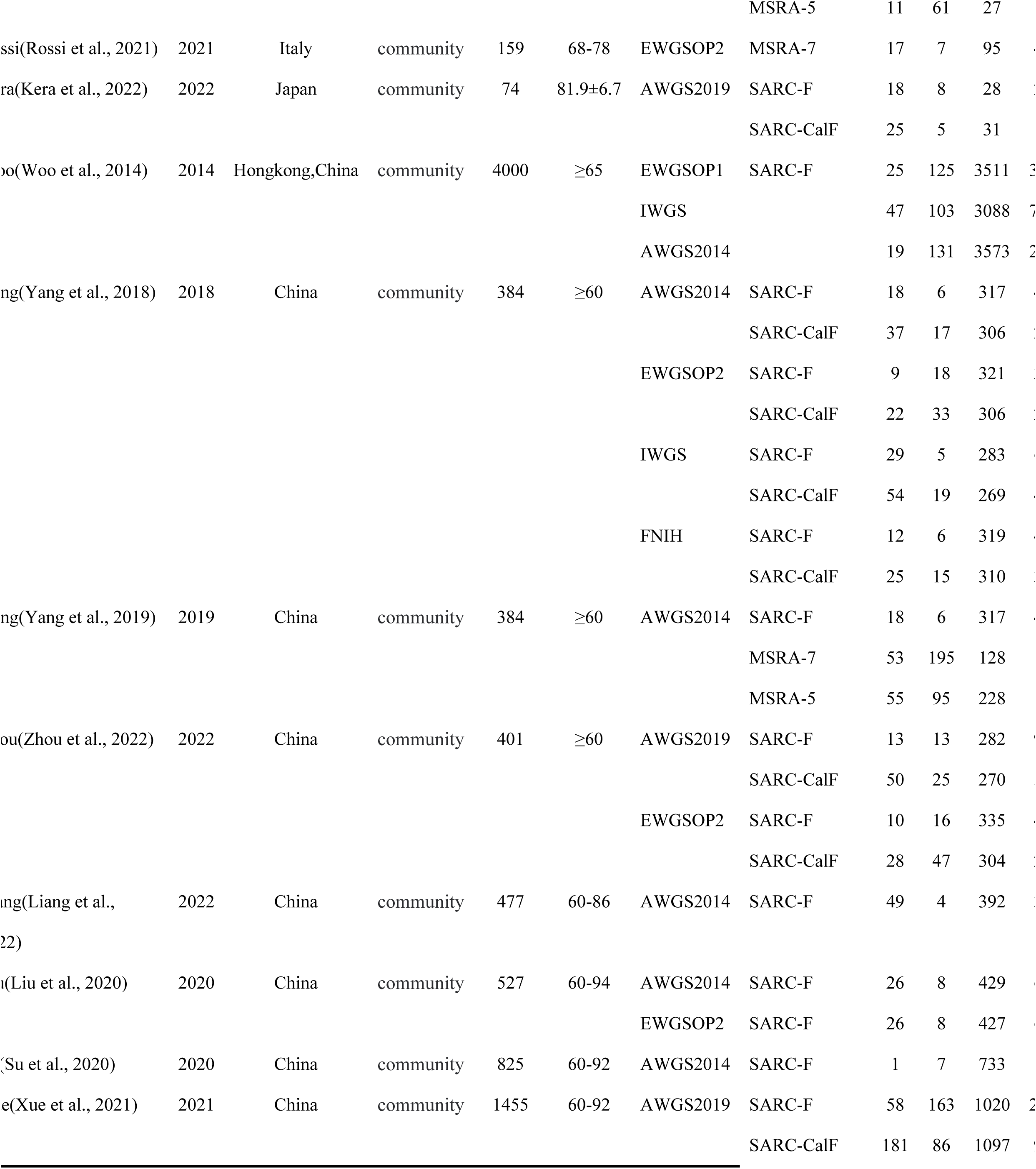
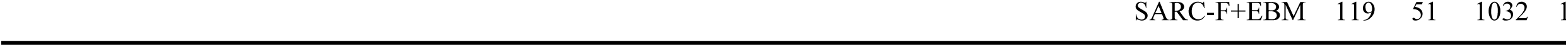
Characteristics of the included studies.

| First author | Year | Country | Population | Sample size | Age, y | Reference standard | Diagnostic criteria | Tp | Fp | Tn | Fn |
| --- | --- | --- | --- | --- | --- | --- | --- | --- | --- | --- | --- |
| Chen(Chen et al., 2020) | 2020 | Taiwan, China | community | 177 | ≥65 | AWGS2019 | SARC-F | 10 | 7 | 78 | 12 |
|  |  |  |  |  |  |  | SARC-CalF | 35 | 20 | 68 | 32 |
|  |  |  |  |  |  |  | MSRA-5 | 55 | 39 | 47 | 53 |
| Erbas(Erbas Sacar et al., 2021) | 2021 | Turkey | community | 456 | ≥65 | EWGSOP2 | SARC-F | 23 | 47 | 351 | 10 |
| Ito(Ito et al., 2021) | 2021 | Japan | community | 139 | ≥65 | AWGS2014 | SARC-F | 2 | 10 | 114 | 5 |
|  |  |  |  |  |  |  | SARC-CalF | 10 | 24 | 100 | 19 |
|  |  |  |  |  |  | AWGS2019 | SARC-F | 2 | 10 | 114 | 5 |
|  |  |  |  |  |  |  | SARC-CalF | 10 | 23 | 101 | 19 |
| Kera(Kera et al., 2020) | 2020 | Japan | community | 1060 | 65-84 | AWGS2014 | SARC-F | 5 | 25 | 906 | 10 |
| Li(Li et al., 2020) | 2020 | China | community | 1009 | ≥60 | AWGS2019 | SARC-F+EBM | 20 | 110 | 812 | 7 |
|  |  |  |  |  |  |  | SARC-F | 36 | 130 | 792 | 17 |
| Lim(Lim et al., 2020) | 2020 | Singapore | community | 230 | 67.2±7.4 | AWGS2019 | SARC-CalF | 27 | 48 | 120 | 3 |
|  |  |  |  |  |  |  | SARC-F | 34 | 36 | 132 | 3 |
| Kim(Kim and Won, 2020) | 2020 | Korea | community | 2123 | 70-84 | AWGS2019 | SARC-F | 23 | 137 | 1841 | 10 |
| Mo(Mo et al., 2021) | 2021 | China | community | 1050 | ≥60 | AWGS2019 | SARC-CalF | 214 | 181 | 606 | 4 |
|  |  |  |  |  |  |  | SARC-F | 125 | 63 | 724 | 10 |

|  |  |  |  |  |  |  |  |  |  |  |
| --- | --- | --- | --- | --- | --- | --- | --- | --- | --- | --- |
| Queiroz(Queiroz et al., 2023) | 2023 | Brazil | community | 258 | 70±8 | EWGSOP2 | SARC-CalF | 10 | 36 | 167 |
|  |  |  |  |  |  |  | SARC-F | 15 | 17 | 186 |
| Piotrowicz(Piotrowicz et al., 2021) | 2021 | Poland | community | 73 | ≥65 | EWGSOP2 | SARC-F | 6 | 8 | 48 |
| Krzywińska(Krzywińska-Siemaszko et al., 2020) | 2020 | Poland | community | 115 | ≥65 | EWGSOP1 | SARC-F | 6 | 14 | 81 |
|  |  |  |  |  |  |  | SARC-CalF | 12 | 11 | 84 |
|  |  |  |  |  |  |  | SARC-F+EBM | 11 | 27 | 68 |
|  |  |  |  |  |  | EWGSOP2 | SARC-F | 6 | 15 | 84 |
|  |  |  |  |  |  |  | SARC-CalF | 10 | 13 | 86 |
|  |  |  |  |  |  |  | SARC-F+EBM | 8 | 29 | 70 |
| Krzywińska(Krzywińska-Siemaszko et al., 2020) | 2020 | Poland | community | 100 | 65-93 | EWGSOP1 | MSRA-7 | 7 | 10 | 70 |
|  |  |  |  |  |  |  | SARC-F | 12 | 8 | 72 |
|  |  |  |  |  |  |  | SARC-CalF | 14 | 42 | 38 |
|  |  |  |  |  |  |  | MSRA-5 | 16 | 56 | 24 |
|  |  |  |  |  |  | EWGSOP2 | MSRA-7 | 7 | 10 | 73 |
|  |  |  |  |  |  |  | SARC-F | 11 | 9 | 74 |
|  |  |  |  |  |  |  | SARC-CalF | 11 | 45 | 38 |
|  |  |  |  |  |  |  | MSRA-5 | 13 | 59 | 24 |
|  |  |  |  |  |  | AWGS2014 | MSRA-7 | 6 | 11 | 73 |
|  |  |  |  |  |  |  | SARC-F | 10 | 10 | 74 |
|  |  |  |  |  |  |  | SARC-CalF | 13 | 43 | 41 |
|  |  |  |  |  |  |  | MSRA-5 | 14 | 58 | 26 |
|  |  |  |  |  |  | FNIH | MSRA-7 | 9 | 8 | 82 |
|  |  |  |  |  |  |  | SARC-F | 2 | 18 | 72 |
|  |  |  |  |  |  |  | SARC-CalF | 9 | 47 | 43 |
|  |  |  |  |  |  |  | MSRA-5 | 9 | 63 | 27 |
|  |  |  |  |  |  | IWGS | MSRA-7 | 6 | 11 | 77 |
|  |  |  |  |  |  |  | SARC-F | 8 | 12 | 76 |
|  |  |  |  |  |  |  | SARC-CalF | 10 | 46 | 42 |
|  |  |  |  |  |  |  | MSRA-5 | 11 | 61 | 27 |  |
| ssi(Rossi et al., 2021) | 2021 | Italy | community | 159 | 68-78 | EWGSOP2 | MSRA-7 | 17 | 7 | 95 | 4 |
| ra(Kera et al., 2022) | 2022 | Japan | community | 74 | 81.9±6.7 | AWGS2019 | SARC-F | 18 | 8 | 28 | 1 |
|  |  |  |  |  |  |  | SARC-CalF | 25 | 5 | 31 |  |
| o(Woo et al., 2014) | 2014 | Hongkong,China | community | 4000 | ≥65 | EWGSOP1 | SARC-F | 25 | 125 | 3511 | 3 |
|  |  |  |  |  |  | IWGS |  | 47 | 103 | 3088 | 7 |
|  |  |  |  |  |  | AWGS2014 |  | 19 | 131 | 3573 | 2 |
| ng(Yang et al., 2018) | 2018 | China | community | 384 | ≥60 | AWGS2014 | SARC-F | 18 | 6 | 317 | 4 |
|  |  |  |  |  |  |  | SARC-CalF | 37 | 17 | 306 | 1 |
|  |  |  |  |  |  | EWGSOP2 | SARC-F | 9 | 18 | 321 | 1 |
|  |  |  |  |  |  |  | SARC-CalF | 22 | 33 | 306 | 1 |
|  |  |  |  |  |  | IWGS | SARC-F | 29 | 5 | 283 | 6 |
|  |  |  |  |  |  |  | SARC-CalF | 54 | 19 | 269 | 4 |
|  |  |  |  |  |  | FNIH | SARC-F | 12 | 6 | 319 | 4 |
|  |  |  |  |  |  |  | SARC-CalF | 25 | 15 | 310 |  |
| ng(Yang et al., 2019) | 2019 | China | community | 384 | ≥60 | AWGS2014 | SARC-F | 18 | 6 | 317 | 4 |
|  |  |  |  |  |  |  | MSRA-7 | 53 | 195 | 128 |  |
|  |  |  |  |  |  |  | MSRA-5 | 55 | 95 | 228 |  |
| ou(Zhou et al., 2022) | 2022 | China | community | 401 | ≥60 | AWGS2019 | SARC-F | 13 | 13 | 282 | 9 |
|  |  |  |  |  |  |  | SARC-CalF | 50 | 25 | 270 | 1 |
|  |  |  |  |  |  | EWGSOP2 | SARC-F | 10 | 16 | 335 | 4 |
|  |  |  |  |  |  |  | SARC-CalF | 28 | 47 | 304 | 1 |
| ng(Liang et al., 2022) | 2022 | China | community | 477 | 60-86 | AWGS2014 | SARC-F | 49 | 4 | 392 | 1 |
| u(Liu et al., 2020) | 2020 | China | community | 527 | 60-94 | AWGS2014 | SARC-F | 26 | 8 | 429 | 6 |
|  |  |  |  |  |  | EWGSOP2 | SARC-F | 26 | 8 | 427 | 6 |
| (Su et al., 2020) | 2020 | China | community | 825 | 60-92 | AWGS2014 | SARC-F | 1 | 7 | 733 |  |
| e(Xue et al., 2021) | 2021 | China | community | 1455 | 60-92 | AWGS2019 | SARC-F | 58 | 163 | 1020 | 2 |
|  |  |  |  |  |  |  | SARC-CalF | 181 | 86 | 1097 | 9 |

### 3.3. Study Quality

The quality evaluation based on QUADAS-2 criteria revealed that the overall quality of the included studies was low. Only 2(Mo et al., 2021, Rossi et al., 2021) studies had a blind design, 10(Kera et al., 2022, Kim and Won, 2020, Krzymiń ska-Siemaszko et al., 2020, Krzymiń ska-Siemaszko et al., 2020, Li et al., 2020, Piotrowicz et al., 2021, Rossi et al., 2021, Yang et al., 2019, Yang et al., 201 8, Zhou et al., 2022) studies included random or consecutive cases, and 19 studies avoided case-control research designs. 17 studies pre determined thresholds, and 7 studies reported the rate of loss to follow-up. All studies provided comprehensive descriptions of baseline information such as gender and age, ensuring consistency and comparability.

### 3.4. Results of traditional meta-analysis

The results of the traditional meta-analysis focused on the pooled sensitivity, specificity, diagnostic odds ratio (DOR), and area under the curve (AUC) of five sarcopenia screening tools. The sensitivities of SARC-F, SARC-CalF, MSRA-5, MSRA-7 were 0.25, 0.59, 0.43, 0.82, and 0.51, respectively. The specificities of these assays were 0.94, 0.82, 0.81, 0.39, and 0.85, additionally, with corresponding DORS of 5, 7, 3, 3 and 6. The AUCs of SARC-F, SARC-CalF, MSRA-5, and MSRA-7 were 0.80, 0.76, 0.70, 0.68, and 0.75. The details are shown in Table 2.

**TABLE 2.** Summary sensitivity, specificity, DOR and AUC of each screening tool.

| Tools | Summary<br>sensitivity(95%CI) | Summary<br>specificity(95%CI) | Summary<br>DOR(95%CI) | Summary<br>AUC(95%CI) |
| --- | --- | --- | --- | --- |
| SARC-F | 0.25(0.19,0.34) | 0.94(0.92,0.96) | 5(4,8) | 0.80(0.76,0.83) |
| SARC-CalF | 0.59(0.59,0.67) | 0.82(0.75,0.88) | 7(4,10) | 0.76(0.72,0.79) |
| SARC-F+EBM | 0.43(0.29,0.59) | 0.81(0.72,0.88) | 3(2,6) | 0.70(0.66,0.74) |
| MSRA-5 | 0.82(0.71,0.90) | 0.39(0.28,0.52) | 3(1,7) | 0.68(0.64,0.72) |
| MSRA-7 | 0.51(0.34,0.67) | 0.85(0.73,0.92) | 6(4,9) | 0.75(0.71,0.79) |

### 3.5. Results of network meta-analysis

#### 3.5.1 Mesh Relationship Chart

Among the studies included in the analysis, SARC-F had the largest sample size, followed by SARC-CalF, MSRA-5, MSRA-7, and SARC-CalF+EBM. The evidence network diagram is depicted in Fig. 2.

**FIGURE 2.**
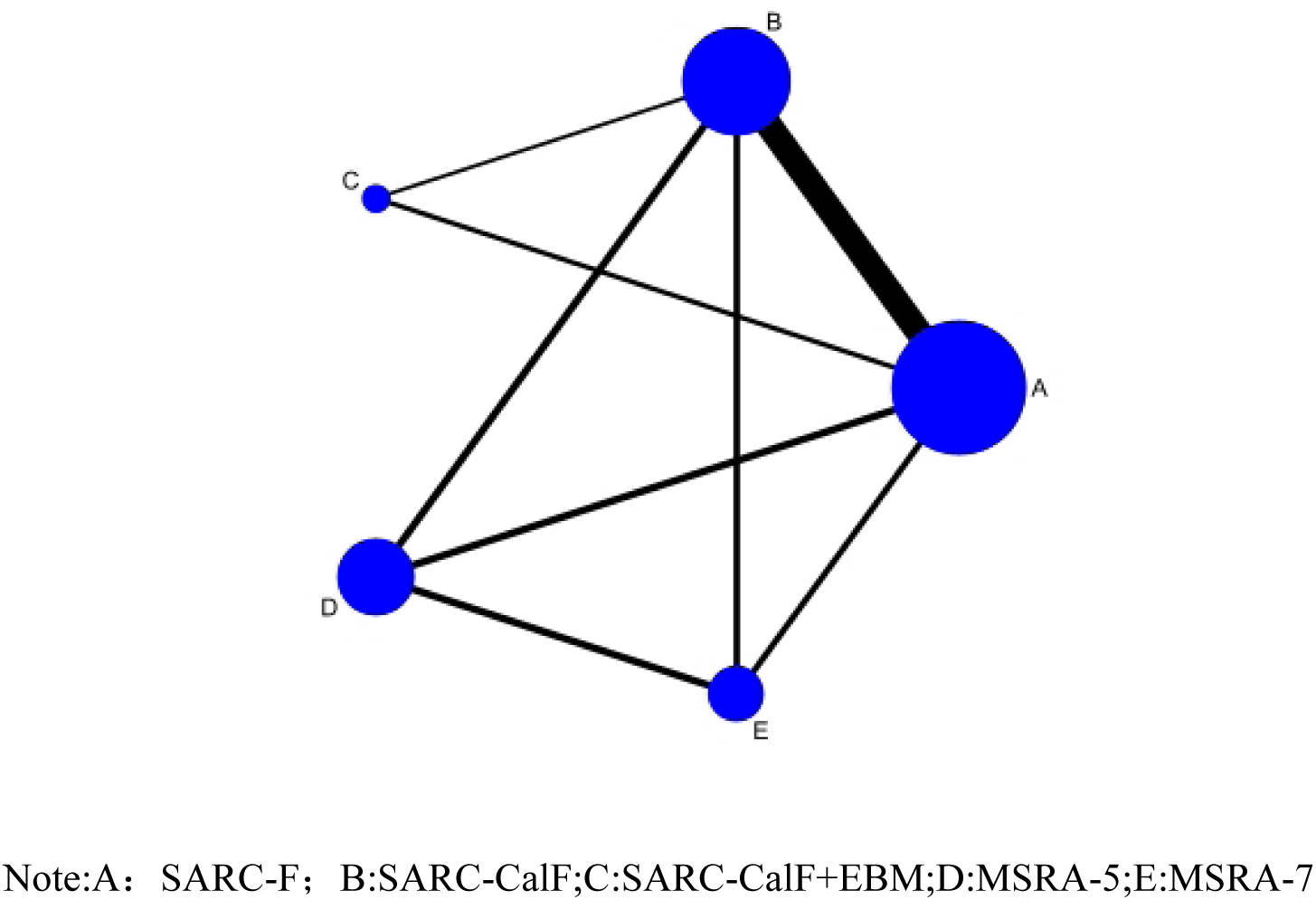
Evidence network diagram.

#### 3.5.2 Consistency test

A consistency test was conducted on each outcome indicator, revealing that there were 9 closed loops, consisting of 4 loops with 3 indicators and 5 loops with 4 indicators. The P values for the direct comparison, indirect comparison, and network comparison result s between different sarcopenia screening tools were all >0.05, indicating a strong level of consistency.

#### 3.5.3 Results of network meta-analysis

The sensitivity results of the network meta-analysis revealed that SARC-F had lower sensitivity compared to SARC-CalF, MSRA-5, and MSRA-7 (P< 0.05). SARC-CalF had lower sensitivity than MSRA-5 and MSRA-7 (P< 0.05). SARC-CalF+EBM had lower sensitivity than MSRA-5 and MSRA-7 (P< 0.05). The specificity results of the network meta-analysis showed that SARC-CalF, SARC-CalF+E BM, MSRA-5, and MSRA-7 had lower specificities than SARC-F, MSRA-5, and MSRA-7 (P< 0.05). The degree was lower for SARC-CalF, SARC-CalF+EBM (P< 0.05), and MSRA-7 was lower than MSRA-5 (P< 0.05). The details are shown in Table 3.

**TABLE 3.**
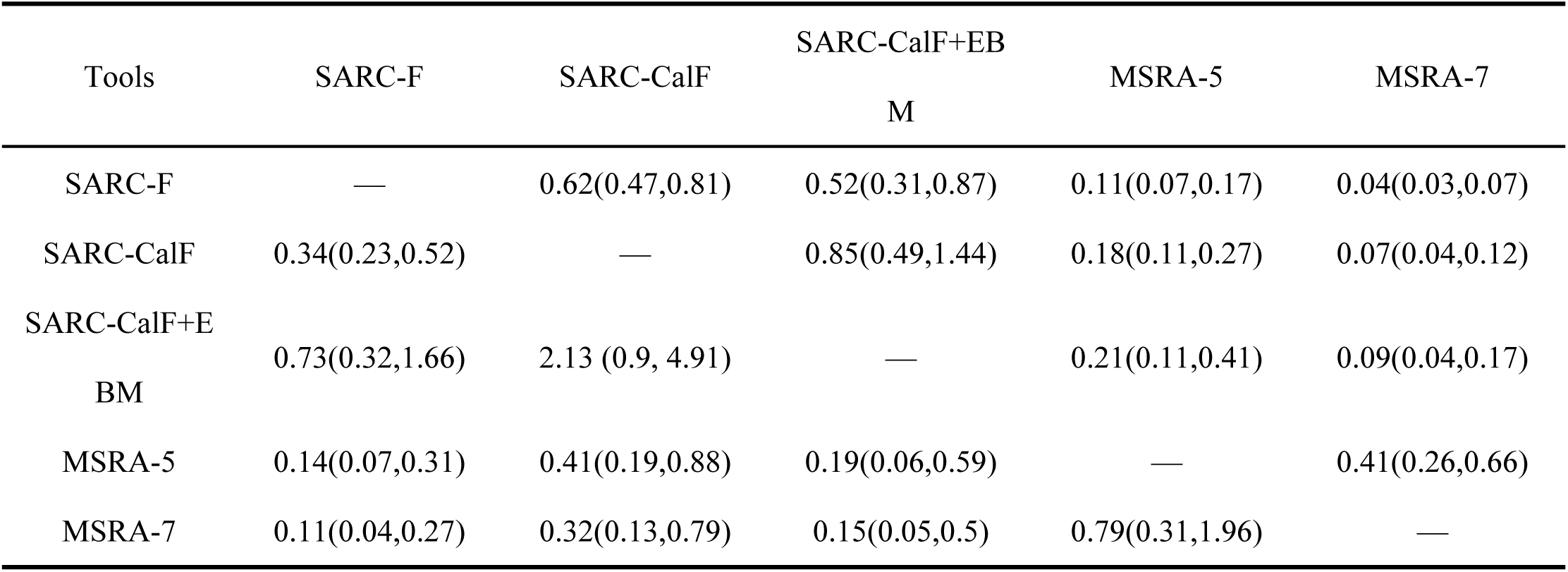
Mesh meta-analysis results of sensitivity (bottom left) an d specificity (top right) of different sarcopenia screening tools [OR (95%CI)]

#### 3.5.4 Sorting of results

The Table 4 provides details on the cumulative ranking probability area under the curve (SUCRA) of various sarcopenia screening tools. It shows that the highest SUCRA values are associated with MSRA-5 sensitivity, SARC-F specificity, and SARC-CalF positive predictive value and negative predictive value. On the other hand, the lowest SUCRA values are observed for SARC-F sensitivity and negative predictive value, as well as MSRA-7 specificity and positive predictive value.

**TABLE 4.** SUCRA values of different sarcopenia screening tools.

| Tools | Sensitivity | Specificity | Positive<br>predictive value | Negative<br>predictive value |
| --- | --- | --- | --- | --- |
| SARC-F | 5.52 (5) | 99.81 (1) | 75.39 (2) | 22.18 (5) |
| SARC-CalF | 49.44 (3) | 68.26 (2) | 93.87 (1) | 83.71 (1) |
| SARC-CalF+EBM | 20.59 (4) | 56.93 (3) | 50.96 (3) | 25.35 (5) |
| MSRA-5 | 92.27 (1) | 24.99 (4) | 26.79 (4) | 79.13 (2) |
| MSRA-7 | 82.17 (2) | 0 (5) | 2.97 (5) | 39.62 (3) |

### 3.6. Publication bias

Comparison-corrected funnel plots were used to assess publication bias by analyzing sensitivity and specificity. The results indicate that the funnel plot is generally symmetrical, although a few studies fall outside the plot. This suggests the possibility of a small sample effect or publication bias in the included literature. See Fig. 3-4 for more details.

**FIGURE. 3.**
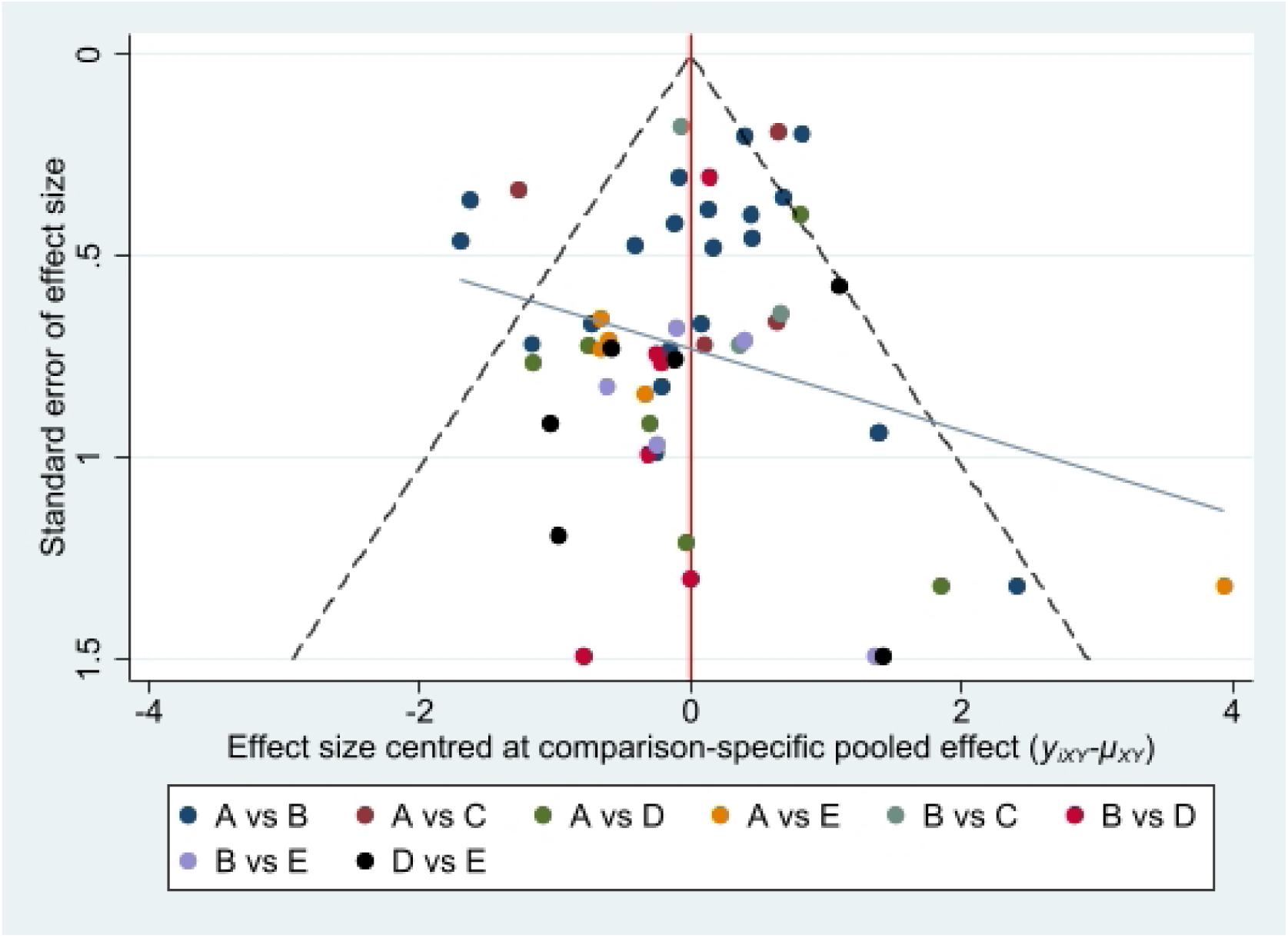
Sensitivity funnel diagram.

**FIGURE. 4.**
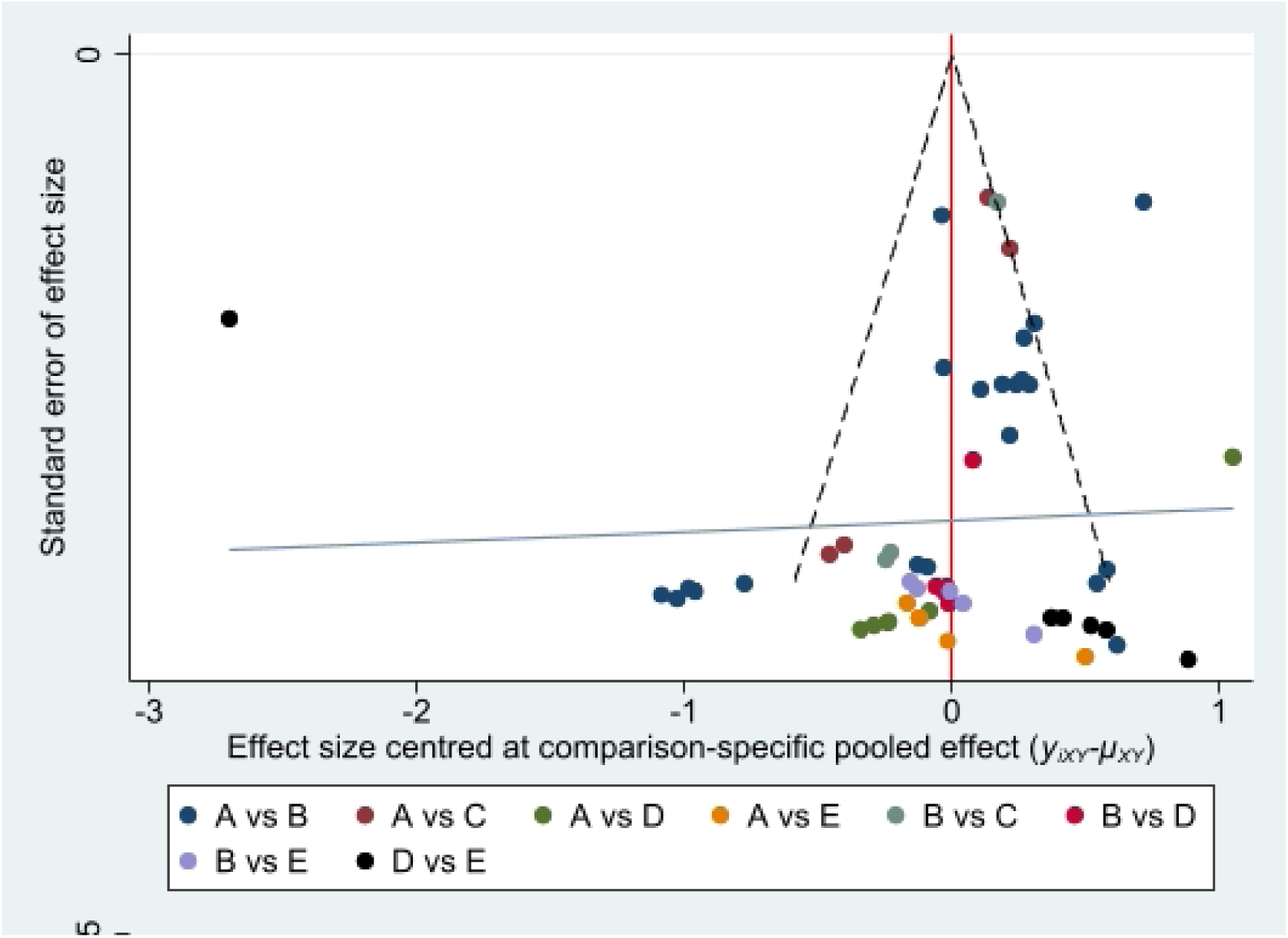
Specificity funnel diagram.

## 4. Discussion

We conducted a systematic review, meta-analysis, and network meta-analysis of 5 common sarcopenia screening tools in 22 studies involving 15,493 community-dwelling older adults and found that M SRA-5 and SARC-F had the highest diagnostic test sensitivity and Specificity. The sensitivity of MSRA-5 in network meta-analysis was found to be consistent with the values reported in a routine meta-analysis of 7 studies. Similarly, the specificity of SARC-F in network meta-analysis was consistent with the values reported in a routine meta-analysis of 20 studies. This implies that MSRA-5 is the most sensitive tool for screening sarcopenia, while SARC-F is the most specific tool for sarcopenia screening.

MSRA, a sarcopenia screening tool proposed by Italian scholar Rossi (Rossi et al., 2017) in 2017, includes age, physical activity level, hospitalization, weight loss, number of daily meals, dairy consumption, and protein consumption. The sensitivity of the MSRA-7 scale is 80.4% with a specificity of 50.1%. However, after excluding the number of daily meals and dairy product consumption, the sensitivity of the MSRA-5 scale remains at 80.4% with a specificity of 60.4%. Furthermore, in 2018, domestic scholar Yang (Yang et al., 2 018) concluded that the Chinese version of China mini sarcopenia risk assessment (C-MSRA) can effectively screen sarcopenia in elderly Chinese communities using the Chinese-language MSRA scale. When comparing C-MSRA-7 and C-MSRA-5, it was found that C-MSRA-5 is more suitable for sarcopenia screening. The reason for this analysis could be that the original MSRA-7 scale items were primarily designed for consumption of dairy products in Western countries, and the nutrition-related items may not be suitable for other countries. As a result, the MSRA-5, after correcting for this factor, has a wider range of applicability and higher screening sensitivity. How ever, the current application of MSRA in screening sarcopenia among community-dwelling elderly individuals is still relatively limited. Further verification is necessary by expanding the sample size.

The SARC-F scale was introduced by Malmstrom (Malmstrom and Morley, 2013) and colleagues in 2013. It primarily relies on patients’ self-reported characteristics of sarcopenia, such as their perception of muscle strength and walking ability. The results may be influenced by the subjective life attitudes and psychological factors of the elderly, resulting in higher specificity but lower sensitivity. Currently, the SARC-F scale is widely recognized and utilized as a simple, safe, and cost-effective tool for screening sarcopenia. Huang et a l. (Huang and Wang, 2020), Chinese scholars, have translated the scale into Chinese for community-dwelling elderly individuals aged ≥6 0 years. Their findings demonstrate that the Chinese version of the SARC-F scale exhibits good reliability and validity, with higher specificity than sensitivity. They suggest that the SARC-F scale can serve as an initial step in community screening for sarcopenia. Gasparik et al. (Gasparik et al., 2020) translated SARC-F into the Romanian version and verified it on 80 elderly people from nursing homes and hospitals. The Cronbach’s alpha coefficient was 0.75, the sensitivity was 0.694, and the specificity was 0.840. Germany’s Drey et a l. (Drey et al., 2020) applied the German version of SARC-F to con duct a test on 117 community outpatients. The results showed that the sensitivity of the scale was 0.75 and the specificity was 0.67. The difference in sensitivity and specificity results between the two studies may be caused by differences in the ratio of male to female subjects and the source of the groups. Due to the low sensitivity of SARC-F, Voelker et al. (Voelker et al., 2021) do not recommend this questionnaire as a screening tool for sarcopenia.

Sarcopenia in community-dwelling elderly individuals is influenced by multiple factors. Prevention should be prioritized over treatment, and accurate and effective screening and assessment are essential for prevention. This study reveals that the MSRA-5 screening tool, although highly sensitive, is rarely used among the elderly in the community. On the other hand, the SARC-F scale primarily focuses on muscle function rather than muscle mass, resulting in high specificity but poor sensitivity. It may miss some individuals at risk. The SARC-CalF scale, which improves the accuracy of the SARC-F scale, requires the measurement of calf circumference (CC). The cut-off point threshold for predicting muscle mass and calf circumference in the SARC-CalF scale (34cm for men and 33cm for women) is determined based on research conducted on Brazilian community-dwelling elderly individuals. However, studies have shown that the cut-off point of calf circumference can be influenced by factors such as gender, age, race, and environment. Therefore, selecting the appropriate cut-off point threshold for calf circumference is crucial in sarcopenia screening. If the cut-off value is set too low and does not account for gender differences, the SARC-CalF scale may result in a lower prevalence of sarcopenia. When nursing staff conduct community screening for sarcopenia in the elderly, they should develop a screening plan that takes into consideration the characteristics of the SARC-F scale, SARC-CalF scale, and MSRA-5 questionnaire, as well as the specific population being screened. In conclusion, this study suggests that an individualized joint screening program should be considered during community sarcopenia screening to minimize false positives and false negatives, thereby improving the accuracy of screening and providing a foundation for the prevention and treatment of sarcopenia.

## 5. Limitations

This study has several limitations:(i) It excludes studies on secondary sarcopenia, which reduces heterogeneity to some extent, but also limits the universality and comprehensiveness of the evidence. (ii) Some screening tools have limited studies, weak support, and a certain degree of bias. (iii) The study only includes Chinese and English literature, which may introduce a certain degree of language bias.

## 6. Conclusion

This study is the first to utilize Bayesian network meta-analysis to compare the diagnostic efficacy of five commonly used community sarcopenia screening tools in the screening of sarcopenia among elderly individuals living in the community. The findings indicate that the MSRA-5 questionnaire exhibits higher sensitivity compared to SARC, while the SARC-F scale demonstrates high specificity. Utilizing multiple scales for screening can potentially decrease the rate of false positive and false negative results. However, further research involving larger sample sizes, multi-center studies, and high-quality clinical trials is necessary to validate these conclusions.

## Data Availability

All relevant data are within the manuscript and its Supporting Information files.

